# Educational Survey of Bidirectional Physician Exchange between St. Damien Pediatric Hospital in Haiti and US Medical Centers

**DOI:** 10.1101/2024.11.26.24317995

**Authors:** Jessica Jordan, Theony Deshommes, Renee Alce, Richard Tucker, Michael P. Koster, Beatrice E. Lechner

## Abstract

**Objective:** To evaluate Haitian and U.S. resident experience with the bidirectional resident physician exchange program between St. Damien Pediatric Hospital in Haiti and partner U.S. medical centers.

**Methods:** A cross-sectional study was carried out among 23 Haitian residents and 51 U.S. residents who participated in the bidirectional resident physician clinical rotation exchange program since its inception. Unique electronic surveys were created and distributed to each group, and quantitative and qualitative analysis of responses was performed.

**Results:** Thirty one responses were obtained; 14 Haitian residents and 17 U.S. residents responded. Several significant differences emerged between the Haitian residents’ and U.S. residents’ experiences. These included motivation for participating in the exchange, perspectives on the most beneficial learning components of the exchange, perceived challenges, and experience with teaching methodology and feedback. Specifically, Haitian residents were more likely to encounter a lack of clinical hands-on experiences than U.S. residents and U.S. residents were more likely to experience language related communication challenges.

**Conclusion:** This survey demonstrates that the bidirectional resident physician clinical rotation exchange experience is a useful vehicle for collaborative learning between residency programs in high- and low-resource settings which benefit both groups, and that the experience is different in key ways for each group. These observations suggest that incorporation of these findings into bilateral exchange programs will strengthen the residency educational partnership between hospitals in low- and high-resource settings, resulting in better trained physicians in both settings.

**Lay Summary:** Haiti is the lowest-income country in the Western Hemisphere with the highest rates of infant and maternal mortality, malnutrition and persons living with HIV/AIDS. The shortage of healthcare professionals, specifically pediatric trained physicians, is a root cause of health disparities in Haiti. The bidirectional residency exchange program between St. Damien Pediatric Hospital in Haiti and its partner US medical centers, which comprise the St. Damien Collaborative, endeavors to help address the pediatrician shortage in Haiti. Through this program, Haitian pediatric residents complete rotations at partner US medical centers, gaining exposure to more technologically advanced subspecialties, while US pediatric residents complete rotations in Haiti, learning about medical cases that are more common in resource-limited settings. This research demonstrated that the exchange experience is different in key ways for Haitian residents visiting US hospitals compared to US residents visiting Haiti, and suggests that the program can be further tailored to reflect these differences in order to strengthen the program, resulting in better trained physicians who will combat the shortage of healthcare professionals in Haiti. These results also provide a roadmap for developing mutually fruitful bidirectional residency exchange partnerships between other high- and low-resource residency training programs.

## Background

Haiti is the poorest country in the Western Hemisphere, with 65% of the population living under the national poverty line, and the highest rates of infant and maternal mortality, malnutrition and persons living with HIV/AIDS.^1,2^ The shortage of healthcare professionals is a root cause of health disparities and it remains a challenge to train an adequate number of physicians in Haiti. For every 10,000 people in Haiti, there are only four health professionals, and trained pediatricians have an even lower ratio.^3^

St. Damien Hospital (Hopital Saint Damien-Nos Petits Freres et Soeurs) is a pediatric hospital that delivers 50,000 essential services per year to Haitian children. After the 2010 earthquake, St. Damien Academic Collaborative was created as a consortium of North American based pediatric hospitals dedicated to building capacity for pediatric care in Haiti.^3,4,5^ In 2013, St. Damien Hospital launched a pediatric residency program with support from St. Damien Collaborative. In a country with only 300 pediatricians and three pediatric residency programs, supporting the St. Damien Hospital-Hopital Bernard Mevs pediatric residency program became the primary mission of St. Damien Collaborative, with a goal of building capacity for pediatric care in Haiti.^6,7^

One component of the residency program is a bidirectional exchange, during which Haitian residents rotate at partner sites in the US and visiting teams from the US rotate in Haiti. Exchange programs offer a unique opportunity for trainees to expand their knowledge and learn from peers in different contexts. Haitian senior residents gain exposure in more technologically advanced subspecialities in the U.S. that are not as well developed in Haiti, and US residents are exposed to cases more common in resource-limited settings, as well as advanced stages of diseases.

Over the past decade, there has been an increase in partnerships between researchers and physicians in lower resourced and higher resourced countries.^8^ One such example is the Pediatric Emergency and Critical Care-Kenya Fellowship training program, built for training pediatricians from Sub-Saharan Africa..^8,10^ However, the St. Damien Collaborative residency exchange program is unique given its bidirectional nature. To our knowledge, pediatric residency programs with a bidirectional exchange do not exist in other low-resource settings.

This study endeavors to describe the experiences of both Haitian and US residents with the bidirectional exchange program, with the aim of strengthening the partnership. Focusing on program strengths and weaknesses provides valuable insight for successful development of combined low-and high-resource setting pediatric training program partnerships aimed at capacity building.

## Methods

Unique electronic surveys were sent to residents who completed rotations at US sites and at St. Damien Hospital in Haiti. Demographic information was collected. In order to assess residents’ experience with clinical feedback, items from the EFFECT questionnaire^12^ were used, a validated measure for clinical teaching.^11,12^ Haitian and US residents were asked general questions about their exchange experience and aspects of the rotation most beneficial for learning how to provide high quality patient care. Haitian residents were asked specifically about their experience with patient-family centered rounds. US residents were asked specifically about exposure to medical cases in Haiti that are not as common in the US. Respondents could choose as many aspects as applied. Haitian and US residents were asked about: challenges experienced during the exchange, specifically language, observer status, cultural challenges and length of rotation; teaching methodology experienced; components of successful partnership observed; quality and content of feedback (residents were presented with statements about feedback and asked to what degree they agreed [strongly disagree/disagree/neither agree nor disagree/agree/strongly agree]); personal support, interactions with clinical teachers; how the exchange helped in their practice of medicine after they returned to Haiti or the US; challenges encountered after their return (potential challenges include changes in goals, shift in attitude towards norms in home country, readjustment to practices in home country, sense of isolation from peers).

Surveys were translated into French for Haitian participants and English surveys were used for US participants. Surveys were distributed as an electronic link via RedCap to residents who completed rotations from 2013-2020. Participants completed a consent form at the beginning of the survey, stating that they understood the survey was completely anonymous. Participant email lists were used and participants were sent three reminder emails to complete the survey. RedCap allowed for anonymous data responses, while tracking individuals who had completed surveys. For quantitative aspects of the survey, mean overall scores were calculated and t-tests were used to test differences in continuous variables and the chi-square test was used to compare categorical responses between US and Haitian residents. Qualitative data were analyzed for themes using standard theme development methods. This project did not meet the federal definition of human subjects research. Therefore, the Brown University IRB declined review.

## Results

### Patient Demographics

14 Haitian residents and 17 US residents responded to the electronic survey. Of the 14 Haitian residents, 12 residents (85.7%) were female, while 12 of 17 US residents (70.6%) were female.

Two Haitian residents graduated from medical school in 2012, 3 in 2013, 6 in 2014 and 3 in 2015. Two US residents graduated from medical school in 2013, 3 in 2014, 2 in 2015 and 3 in 2016. There was no significant difference in year of graduation between Haitian and US residents.

Of survey respondents, one Haitian resident (7%) participated in the exchange program in 2015, 3 (21%) in 2017, 6 (43%) in 2018, 2 (14%) in 2019, and 2 (14%) in 2020. Three US residents (18%) participated in 2013, 2015 and 2019, 1 (6%) participated in 2016, and 5 (29%) in 2017. There was a significant difference in exchange year of Haitian residents and US residents (p=0.041), with more US residents participating in earlier years compared to more Haitian residents in later years.

### Length of Rotation Abroad

The majority of Haitian residents (10, 71%) spent 1-2 months at their rotation site. 4 (29%) Haitian residents reported <1 month at their rotation site. Eight (47%) US residents spent 1-2 months at their rotation site, 7 (41%) spent <1 month, and 1 (6%) spent 2-3 months at their site. There was no significant difference in length of rotation abroad between US and Haitian residents.

### Rotation Sites

St. Damien Collaborative is composed of 15 US sites at which Haitian residents in the program completed rotations, as well as 5 sites at which Haitian respondents rotated (Table 1).

**Table 1.** St. Damien Collaborative Rotation Site.

| **St. Damien Collaborative Rotation Sites: Haitian Residents (n=14)** |  |
| --- | --- |
| 1. Akron Children's Hospital<br>2. University of Minnesota Masonic Children's Hospital<br>3. Hasbro Children's Hospital<br>4. Baystate Children's Hospital<br>5. University of Nebraska Medical Center<br>6. Rainbow Babies and Children's Hospital<br>7. Montreal Children's Hospital<br>8. Children's Hospital of the King's Daughters<br>9. East Tennessee State<br>10. Connecticut Children's Medical Center<br>11. Cohen's Children's Medical Center<br>12. Nationwide Children's Hospital<br>13. Walsh University<br>14. New York Presbyterian<br>15. Morgan Stanley Children's Hospital and Ascension Global Mission |  |
| **Sites at which survey respondents rotated:** |  |
| University of Nebraska Medical Center | 2 (14%) |
| Rainbow Babies and Children's Hospital | 2 (14%) |
| Montreal Children's Hospital | 6 (43%) |
| Children's Hospital of the King's Daughters | 2 (14%) |
| Cohen's Children's Medical Center | 2 (14%) |

### Subspecialty Programs

Haitian residents could engage in multiple subspecialty rotations at US sites. Subspecialties reported were cardiology (57.1%), hospital medicine (35.7%), gastroenterology (28.6%), emergency medicine (14.3%), pediatric critical care (14.3%), hematology/oncology (7.2%), infectious disease (7.1%), neonatology (7.1%), and pediatric neurology (7.1%).

### Interest in Bilateral Exchange Program

Haitian and US residents were asked about their motivation for participating in the bilateral exchange program (Table 2). Personal interest displayed a significant difference, with more US residents reporting personal interest as motivation than Haitian residents (p<0.005).

**Table 2.**
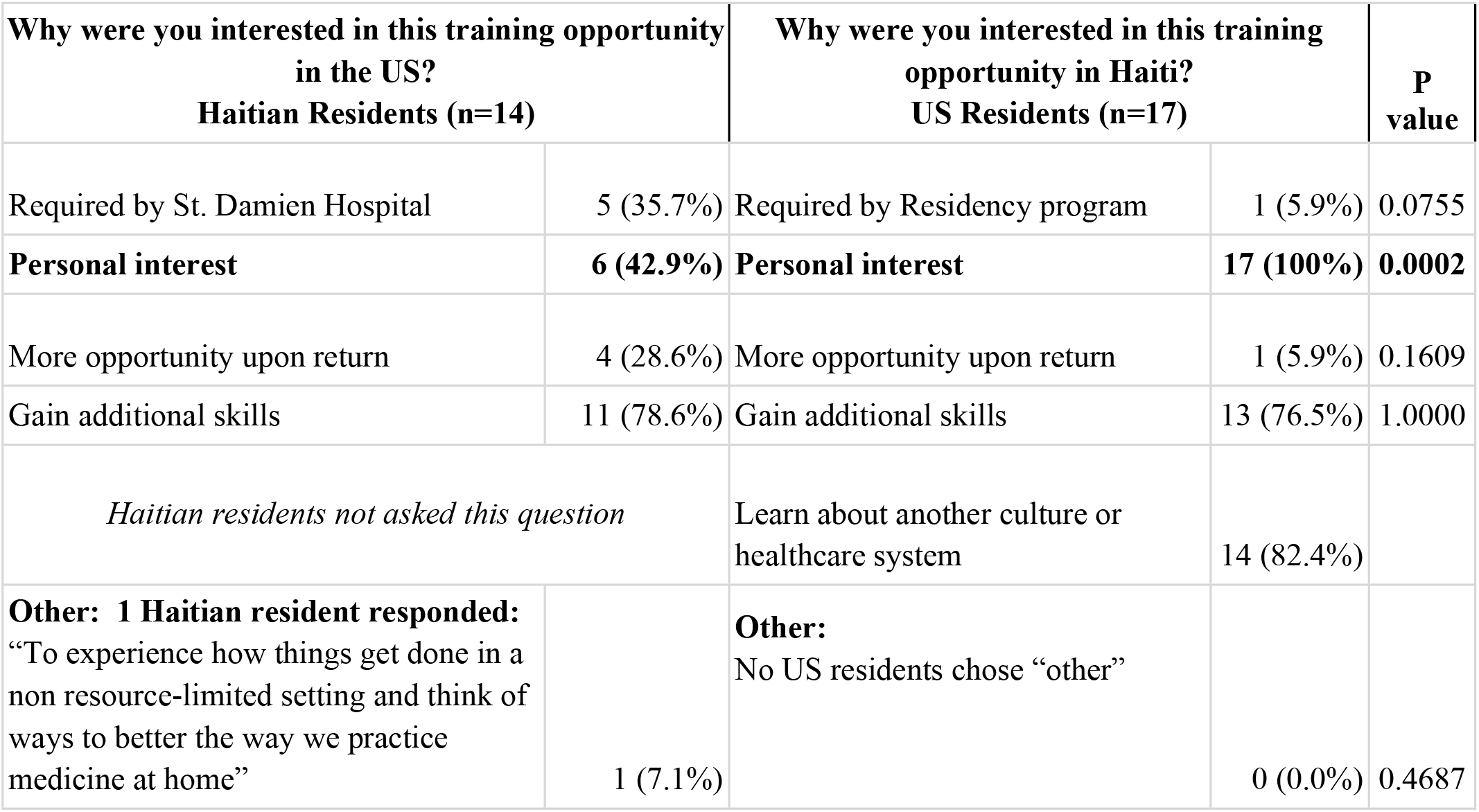
Interest in the bilateral exchange program.

### General Exchange Experience

There were no significant differences in responses between Haitian and US residents regarding general exchange experience. 92.9% Haitian residents and 88.2% US residents reported they received adequate housing support while participating in the exchange. 64.3% Haitians and 88.2% US residents reported the program length was sufficient for learning. 64.3% of Haitian residents and 82.4% of US residents reported they received adequate pre-departure support before participating in the exchange. 64.3% of Haitian residents and 64.7% of US residents responded they received adequate financial support while participating in this exchange. Finally, 50% Haitians and 64.7% US residents reported receiving adequate cultural awareness training while participating in the exchange.

### Learning benefits

There was a significant difference in residents’ perceptions of the benefit of conferences and lectures. Haitian residents valued them more than the US residents (p=0.004). Additionally, Haitian residents valued resident core lectures more than US residents (p=0.015). Finally, there was a trend towards US residents choosing managing patient care as a beneficial aspect in comparison to Haitian residents (p=0.061). There were no other significant differences between US and Haitian residents (Table 3).

**Table 3.** General Experience with the Bidirectional Exchange.

| **What aspects of your rotation were the most beneficial for learning how to better provide high quality patient care?** | **Haitian Residents**<br>**(n=14)** | **US Residents**<br>**(n=17)** | **P value** |
| --- | --- | --- | --- |
| **Conferences and lectures** | **11 (78.6%)** | **3 (17.6%)** | **0.0036** |
| Shadowing in clinic | 8 (57.1%) | 5 (29.4%) | 0.2804 |
| **Resident core lectures** | **5 (35.7%)** | **0 (0%)** | **0.0149** |
| Pairing up with residents to care for patients | 4 (28.6%) | 7 (41.2%) | 0.4719 |
| Examining and interacting with patients independently | 6 (42.9%) | 7 (41.2%) | 1.0000 |
| Managing patient care | 2 (14.3%) | 8 (47.1%) | 0.0605 |
| **Haitian Residents:**<br>Experiencing patient-family centered rounds | 8 (57.1%) |  |  |
| **US Residents:** Exposure to medical cases in Haiti that are not as common in the US |  | 14 (82.4%) |  |
| **Which of the following were challenges during your rotation?** |  |  |  |
| **Language** | **4 (28.6%)** | **11 (64.7%)** | **0.0416** |
| **Observership status** | **8 (57.1%)** | **1 (5.9%)** | **0.0049** |
| Cultural challenges | 5 (35.7%) | 9 (52.9%) | 0.3075 |
| Length of rotation | 4 (28.6%) | 2 (11.8%) | 0.3828 |
| **Which teaching methodologies were you exposed to?** |  |  |  |
| Reviewed learning objectives | 3 (21.4%) | 4 (23.5%) | 1.0000 |
| Asked me to explain my choice for a particular approach. | 9 (64.3%) | 6 (35.3%) | 0.2872 |
| Discussed possible clinical courses and/or complications. | 8 (57.1%) | 8 (47.1%) | 1.0000 |
| **Stimulated me to ask questions.** | **12 (85.7%)** | **5 (29.4%)** | **0.0060** |
| Stimulated me to actively participate in discussions. | 7 (50%) | 7 (41.2%) | 1.0000 |
| Explained complex medical issues clearly. | 6 (42.9%) | 7 (41.2%) | 1.0000 |
| Asked me to give a formal presentation at the end of my rotation. | 5 (35.7%) | 7 (41.2%) | 0.7257 |
| **What components of successful partnership did you observe during the exchange?** |  |  |  |
| Open communication | 12 (85.7%) | 14 (82.4%) | 1.0000 |
| Ability to compromise | 4 (28.6%) | 9 (52.9%) | 0.1657 |
| Mutual respect | 13 (92.8%) | 13 (76.5%) | 0.6586 |
| **Shared vision** | **4 (28.6%)** | **12 (70.6%)** | **0.0320** |

### Challenges during exchange program

US residents felt language was a more significant challenge than Haitian residents (p=0.042). Haitian residents responded that observership status was a significant challenge, compared to US residents (p=0.005) (Table 3).

### Teaching methodology

One statistically significant difference emerged between US and Haitian residents, with more Haitian residents reporting feeling more stimulated to ask questions (p=0.006) during their exchange. Otherwise, Haitian and US residents had similar reported experiences with teaching methodology.

### Components of a successful partnership

More US residents observed a shared vision during their rotation than Haitian residents (p=0.032).

### Feedback

The percentage of residents who agreed (agree/strongly agree) with each statement was presented. More Haitian residents reported they received feedback in a way that was not condescending or insulting in comparison to their US counterparts (p=0.018). Additionally, there was a trend towards more Haitian residents reporting they received feedback that indicates what they were doing correctly in comparison to US residents (p=0.066) (Table 4).

**Table 4.** Experience with feedback during the rotation.

| **Feedback Experience** | **Haitian Residents (n=14)**<br>**% Strongly Agree/Agree** | **US Residents (n=17)**<br>**% Strongly Agree/Agree** | **P value** |
| --- | --- | --- | --- |
| I received feedback based on concrete observations of me | 7 (50%) | 4 (26.7%) | 0.2635 |
| **I received feedback in a way that was not condescending or insulting.** | **11 (78.6%)** | **5 (33.3%)** | **0.0180** |
| **I received feedback that indicates what I am doing correctly.** | **9 (64.3%)** | **4 (26.7%)** | **0.0656** |
| I received feedback that indicates what I can improve. | 8 (57.1%) | 4 (26.7%) | 0.1394 |
| I received feedback on my clinical and technical skills. | 7 (53.8%) | 4 (26.7%) | 0.2458 |
| I did not receive feedback. | 5 (35.7%) | 8 (53.3%) | 0.2817 |

### Personal support

There were no significant differences between Haitian and US residents who agreed (agree/strongly agree) regarding individual support experiences. 92.9% of Haitian residents and 76.5% of US residents agreed that clinical teachers treated them respectfully. 85.7% of Haitian residents and 76.5% of US residents responded that clinical teachers were enthusiastic instructors and supervisors. 92.9% of Haitian residents and 88.2% of US residents agreed that clinical teachers were open to questions. Finally, 85.7% of Haitian residents and 82.4% of US residents felt welcomed by clinical departments, hospital and community.

### Post rotation benefits

The majority of Haitian residents (78.6%) responded that this experience helped broaden their understanding of the US health system. 71.4% of Haitian residents reported they learned skills that will help patient care. 64.3% of Haitian residents responded that this experience helped them understand more about US residency training. 50% of Haitians responded that subspecialty rotations will help further their career and 50% responded that this experience will strengthen their partnership with US institutions in the exchange. 35.7% of Haitian residents reported that material learned in the US will help formulate didactic lectures and teaching conferences back home.

There were no significant differences in responses between two groups. The majority of US residents (94.1%) felt this experience helped them understand the Haitian health system. 58.8% of US residents reported they learned clinical skills that will help patient care and 58.8% reported this experience helped them understand more about Haitian residency training. 58.8% also reported they learned material that will help formulate didactic lectures and teaching conferences back home. 47.1% of US residents felt this experience strengthened the partnership with St. Damien Hospital.

### Post rotation challenges

There were no significant differences in responses between Haitian and US residents. Two challenges displayed a trend toward significance. 64.3% of Haitian residents responded they experienced a change in goals and priorities, while only 29.4% of US residents reported the same experience (p=0.153). US residents (58.8%) reported more difficulty with readjustment to practices at home compared to their Haitian counterparts (35.7%) (p=0.178). Both Haitian (50%) and US (52.9%) residents reported difficulties with the shift in attitude towards norms in their home countries after their return. 5.9% of US residents described a sense of isolation from peers after their return.

### How do subspecialty programs abroad affect residents’ careers?

When asked if subspecialty programs abroad encourage trainees to leave their home country to practice elsewhere, 14.3% of Haitian residents responded yes, 42.9% responded no, and 42.9% responded unsure. When asked how their experience in Haiti might impact the US residents’ careers and perspectives upon return, 88.2% responded that it encouraged them to incorporate global health into their career, 94.1% responded that it changed their perspective on the US healthcare system and 88.2% responded that it directly influenced their decision to work with underserved communities in their current practice.

### How else did the experience help in the practice of medicine after your return?

Haitian residents were asked in an open ended manner how else their experience abroad will help in the practice of medicine after their return. Several themes emerged:

### Theme 1: Value of family centered rounds

Many Haitian residents commented that family centered rounds will help in their practice of medicine after they return to Haiti. Representative quotes include: “This experience particularly shaped the way I interact with patients and their families and helped me communicate better.”

### Theme 2: Better clinical understanding and diagnostic skills

In addition, many Haitian residents stated they developed a better clinical understanding and diagnostic skills. One resident contributed, “this experience helped me to find more different possible diagnostics”. Others stated the experience helped them “advocate for labs”, and develop “better clinical understanding”. One resident felt, “experiencing how things get done in a non resource-limited setting and thinking of ways to better the way we practice medicine at home” was very beneficial. Another resident stated, “I was impressed by the care of patients with chronic disease, most of them would die if they were in Haïti.”

### Theme 3: Develop communication skills and skills for emergency settings

Many Haitian residents commented on how their experience helped them develop other skills, such as “skills to take charge of emergencies” and “communication skills with the medical team.”

### Theme 4: Strengthen ability to see the whole patient

Many residents commented this experience helped them see patients holistically, which is beneficial to their practice in Haiti. One participant stated, “I learned to see the whole patient, not just the pathology.” Another noticed, “US pediatric residents have more time to study and discuss patients. They don’t have as much work as the Haitian residents. We have a lot of cases and very ill patients to manage”, which makes it hard to focus on the whole patient.

### US Residents: How else did the experience abroad help in the practice of medicine after your return?

US residents were also asked in an open ended manner how their experience abroad will help in the practice of medicine after their return. One resident responded, “I focused on much more ultrasound given how useful it was in the global context.” Another stated, “I learned how to practice medicine with less.”

### How did the bidirectional aspect of exchange enhance your experience?

When Haitian residents were asked if the bidirectional aspect of this exchange enhanced their experience, 93% responded yes. Common themes from Haitian responses about benefits of a bidirectional exchange included learning from residents, but also sharing their own experiences, using skills learned in the US to improve care in Haiti, adopting a multidisciplinary approach to managing patients and forming connections with other physicians, with whom they still discuss cases. Representative quotes include “I feel more comfortable managing my patient” and “I strongly believe I learned a lot but also I had something to teach and share from my own experience.” Another Haitian resident stated, “in addition to learning how to better search for a solution, I have kept contacts with whom I continue to discuss certain cases.” Two other residents commented on the shift in perspective of their own country, such that “you see what other countries do… take what is possible for you to improve yourself and the care you are giving to your patients” and “it helped me discover other health care system and realize what improvements are necessary in my country.” One Haitian resident explained “We don’t have any subspecialities in Haiti. The experience overseas makes us want to do one as well.”

### Future improvements in the exchange program

Residents were surveyed on future improvements they would like in the bidirectional exchange program. Several themes emerged, including better support, greater exposure to different hospital services, more direct care and less observation, and better collaboration between US and Haitian residents. Themes and representative quotes from Haitian residents are included in Table 5.

**Table 5.** Haitian Resident Perspective: Future Improvements in the Bilateral Exchange Program.

| **Theme** | **Representative Quotes** |
| --- | --- |
| Better financial, language, and housing support | - "St. Damien does not offer adequate financial resources. We are residents struggling with all our personal and financial problems."<br>- "Clear information on financing required to make the trip in order to avoid bad surprises."<br>- "It would help for Haitian residents who are not fluent in English to get help with translation." |
| Greater exposure to different hospital services | - "To go into other services when we have finished our service because we would like to spend as much time as possible in hospitals."<br>- "We need more training in things that we don't have in Haiti, such as ultrasound and simulations... because we do this [rotation] for patients, not for ourselves." |
| Increased direct care, decreased observation | - "We examine patients with other residents, rather than just an observation internship"<br>- "Sometimes shadowing is difficult. They need to let us talk to patients." |
| Better collaboration between US and Haitian residents | - "Make a pair of residents that would evolve together and exchange about respective cases." |
| Length of rotation | - "One month is insufficient... it will be more beneficial for residents to stay for two months to learn more." |

### Challenges

Haitian residents were asked to comment on any other challenges encountered after their return to Haiti. Challenges included lack of materials available in their home country and having to promote evidence-based medicine to their colleagues, “who were not always open to trying new things or learning from what a resident had to share.”

US residents were also asked to comment on any other challenges encountered after their return to the US. One resident stated they, “went from the PICU in Haiti to the PICU in the US and the difference in access and resources was staggering.”

## Discussion

The bidirectional residency exchange program between St. Damien Pediatric Hospital in Haiti and US medical centers is an example of a sustainable medical collaborative focused on building local capacity in a resource limited setting. In 2018, Sors. et al coined the term “reciprocal innovation” while reflecting on the AMPATH program in Kenya, a 30 year partnership between Indiana University in the US and Moi University in Kenya and Referral Hospital in Kenya and a consortium of North American academic institutions led by IU.^13^ Reciprocal innovation highlights the power of a bidirectional and iterative exchange of ideas, resources and innovations to address health challenges in diverse global settings. It illuminates the importance of reciprocity and equity in bilateral exchanges and partnerships in global health, leading to stronger and longer partnerships. The bidirectional residency exchange program in Haiti and the US is another example of reciprocal innovation. It is essential to program improvement to elicit feedback from program participants. This survey is one of the first to examine experiences of residents participating in a bidirectional exchange between a resource limited and a high-resource medical setting, and to explore their experiences with support, teaching methodology, perceived strengths and weaknesses of the exchange, and challenges.

The survey demonstrates the residency exchange experience is different in key ways for Haitian and US residents, and suggests the partnership can be further tailored to reflect survey feedback. These findings expand upon observations in other settings of bidirectional exchange. For example, Carrillo et. al surveyed international surgeons and their hosts who participated in pediatric clinical observerships in North America and found limited funding was a major challenge faced by both parties.^14^ Furthermore, language and cultural barriers, more robust pre-visit orientation and support, access to observing surgical procedures and increased post-visit feedback were all identified as methods to improve the exchange experience.^14^ Similarly, researchers studying the exchange program between University Teaching Hospitals of Lusaka, Zambia and University of Maryland, Baltimore found funding and resource limitations were major challenges for international rotators in the exchange.^15^ Qualitative data from Haitian residents provided more insight into their experience, highlighting the value of family-centered rounds, clinical, diagnostic and communication skills, emergency management, and the value of seeing the whole patient. These survey responses highlight strengths and weaknesses of the programs in Haiti and the US. Incorporating this crucial information into the bidirectional exchange program will strengthen the program and partnership between Haitian and US members of St. Damien Collaborative, resulting in better trained physicians who will combat the shortage of healthcare professionals in Haiti. Beyond this, our findings highlight the importance of partnering with pediatricians training and working in low-resource settings to build capacity, as well as underscoring differences in experiences and educational and supportive needs of medical trainees from low-resource settings. Thus, this survey shines a light on a pathway to improving bilateral exchange partnerships between low- and high-resource settings for building capacity in low-resource settings.

## Data Availability

Accession numbers and DOIs will be made available after acceptance. There are no legal or ethical concerns or restrictions.

## Funding

This work was supported by Warren Alpert Medical School Scholarly Concentration in Global Health.

## Acknowledgements

Hasbro Children’s Hospital Mona Leroy Scholarship for Haitian Resident Exchange.

## Notes

### Competing Interest Statement

The authors have declared no competing interest.

### Author Declarations

This project did not meet the federal definition of human subjects research. Therefore, the Brown University IRB declined review.

